# Genetic propensity to mental health traits and their associations with social connection phenotypes: evidence from the English Longitudinal Study of Ageing

**DOI:** 10.1101/2024.11.14.24317306

**Authors:** Saoirse Finn, Olesya Ajnakina, Feifei Bu, Daisy Fancourt

**Affiliations:** Department of Behavioural Science & Health, Institute of Epidemiology & Health Care, University College London, 1-19 Torrington Place, London, WC1E 7HB, UK

## Abstract

There is a wealth of phenotypic literature on the interplay between mental health and social connections. However, how genetic propensity to mental health traits may be associated with distinct social connections phenotypes (i.e., structural, functional and quality aspects) is largely unexplored. Using polygenic scores (PGSs), we explored the associations between genetic propensity for five mental health traits (**PGS**_**depressive-symptoms**,_ **PGS**_**anxiety**,_ **PGS**_**bipolar- disorder**,_ **PGS**_**schizophrenia**,_ **PGS**_**wellbeing**_) and four social connection phenotypes (social isolation, loneliness, social support and relationship strain). Linear regressions were conducted in a representative sample of unrelated older adults living in the UK, and analyses were controlled for age, sex, and principal components to account for population stratification. The results show that higher **PGS**_**depressive-symptoms**_ was associated with greater loneliness (B=0.11, CI-95%=0.07, 0.15) and relationship strain (B=0.09, CI-95%=0.05, 0.13) and lower social support (B=-0.07, CI-95%=-0.13, -0.01). Higher PGS_anxiety_ was associated with higher social isolation (B=0.05, CI-95%=0.00, 0.10) and greater relationship strain (B=0.05, CI-95%=0.01, 0.09). Higher **PGS**_**bipolar-disorder**_ was associated with greater loneliness (B=0.05, CI-95%=0.01, 0.09) and relationship strain (B=0.06, CI-95%=0.02, 0.10). Higher **PGS**_**wellbeing**_ was associated with lower loneliness (B=-0.07, CI-95%=-0.11, -0.03), relationship strain (B=- 0.07, CI-95%=-0.11, -0.03), and social isolation (B=-0.07, CI-95%=-0.12, -0.02), and greater social support (B=0.14, CI-95%=0.08, 0.21). This suggests differential associations between different mental health PGSs and distinct aspects of social connections, indicating a nuanced picture. Our findings confirm that genetics play a role in having adequate social connections, which can be supported through social, community, and cultural schemes. They also highlight that genetic confounding is important when using observational data assessing the associations between mental health and social connections.

## Introduction

Social connections (interchangeably referred to as ‘social relationships’ or ‘social connectedness’) relate broadly to the social interactions and links between individuals or groups. The dominant conceptualisation of social connections categorises them into structural, functional and quality aspects (Holt□ Lunstad, 2024). Structural aspects refer to the presence and linkages among social relationships and roles (e.g., social network size and amount of social contact). Functional aspects are related to the functions given by or perceived from one’s connections (e.g., loneliness, social support), and quality aspects refer to whether these connections are positive or negative (e.g., relationship strain).

Social connections and mental health are bidirectionally interrelated (Hsueh et al., 2019; Turner et al., 2022). Lower levels of mental health (e.g., depression) are associated with higher loneliness (Cohen-Mansfield et al., 2016; Dahlberg et al., 2021), social isolation (Hakulinen et al., 2016; Wen et al., 2023), and relationship strain (Ali et al., 2022; Hakulinen et al., 2016), as well as lower social support (Hakulinen et al., 2016; Lee & Tan, 2019). In reverse, social connections, such as loneliness, can act as antecedents to mental ill health (Lim et al., 2020). However, this research has largely focused on phenotypic data: self-reported data on social connections and mental health (via validated or commonly used scales). We know very little about how genetic propensity to mental health affects social connections. This is highly relevant given that some individuals may have predispositions towards mental health traits that might not be fully expressed phenotypically or that fall below clinical diagnostic thresholds but may, nonetheless, influence social behaviours (Anttila et al., 2018; Ware et al., 2017).

In recent years, there has been promising novel genetics research on mental health and social connections using polygenic scores (PGSs), which have been made possible by advances in genome-wide association studies (Abdellaoui et al., 2023; Visscher et al., 2017). PGSs provide a single-value genetic estimate of a phenotype (i.e., an individual’s genetic propensity to a trait) by summarising the estimated effect of many genetic variants, which have been shown to capture a far greater share of the variance of a trait than any specific genetic variant on its own (Dudbridge, 2013). Several studies have laid the groundwork for exploring how mental health PGSs are associated with social connection phenotypes. For example, a higher genetic propensity to depressive symptoms, major depressive disorder (MDD), bipolar disorder and schizophrenia (SCZ) have been found to be associated with higher loneliness (Abdellaoui et al., 2018). A higher genetic propensity to SCZ has also been associated with lower relationship satisfaction (Socrates et al., 2021). In addition to mental health symptomatology, a higher genetic propensity to wellbeing has also been associated with lower loneliness and greater relationship satisfaction (Richardson et al., 2019). Overall, these analyses involving PGSs and social connections have provided important insight by showing associations between genetic propensity for psychological traits and social phenotypes.

However, there are several limitations to existing research. Most research on genetic propensity to mental health traits and associations with social connection phenotypes in unrelated samples have used outcome-wide approaches involving multiple different phenotypes (i.e., personality, physical health, mental health, and health behaviours). Whilst this has allowed for the identification of promising relationships, there are several significant consequences. First, analyses have been undertaken outside theoretical frameworks (e.g., combining loneliness and social isolation in the same measure despite the two being unique constructs). Second, some social connection phenotypes have been assessed repeatedly as outcomes (i.e., loneliness) while others have had less attention (i.e., social isolation, social support and relationship strain). Third, given the breadth of these studies, discussions of the findings relating to social connections have often been limited or nonexistent. If we are to truly understand the meaning and relevance of these associations for human behaviour, there is a need to integrate work on genetics further with social science in order to capitalise on the theoretical knowledge of complex social constructs within genetic analyses (Harden & Koellinger, 2020). Additionally, despite the uncertainty of the exact mechanisms underpinning such associations in unrelated population samples, utilising PGSs in observational studies can provide some insight into the temporality between mental health and social connections, help us pinpoint any genetic confounding for future studies, and allow subthreshold genetic propensity to mental health traits to be explored where phenotypic data is not available (i.e., assessing genetic propensity to SCZ in non-clinical general population samples).

Therefore, to advance our understanding of the associations between genetic propensity for mental health traits and social connections phenotypes, we took two novel approaches: using data from a large national cohort study of unrelated older adults living in England and conceptualising our social connections phenotypes within social theory. We considered multiple PGSs of mental health, mental illness and wellbeing alongside social connection phenotypes relating to structural, functional and quality aspects of social connections.

## Methods

### Sample

We analysed data from the English Longitudinal Study of Ageing (ELSA); an ongoing nationally representative study of adults over the age of 50 living in England which started in 2002/2003 (Steptoe et al., 2013). Blood sampling for DNA extraction and storage was collected at Wave 2 (2004/2005), refreshed at Wave 4 (2008/2009), and genotyped in 2013- 14 (Ajnakina & Steptoe, 2022). In ELSA, PGSs are available for 7,182 individuals of White European ancestry. We focused on ELSA wave 2 data, which involves n=9,432 individuals, for whom genetic data was available for n=5,824. We excluded individuals who were younger than 50 years of age and we restricted the final analytical sample to complete cases across all PGSs, outcome variables and covariates in order to compare models, providing a final analytic sample of n=5,257. We only included individuals of White European ancestry due to the available genetic data; the disparities of this are outlined in the discussion. For blood sampling and genotyping details see Supplementary A.

#### Deriving polygenic scores

To create PGSs, SNP weights from GWAS or GWAS meta-analyses of the trait under consideration were used from the most up-to date studies at the time of derivation (2018- 2022) (Ajnakina & Steptoe, 2022). These GWASs did not include ELSA data, or on request, removed ELSA data and reran analyses. The PGSs were derived using the same validated statistical methods as the Health and Retirement Study (HRS) (E. Ware et al., 2021; E. B. Ware et al., 2017). PGSs were constructed using GWAS SNP results significant at a p-value threshold of 1. SNPs were all directly genotyped, and none were imputed due to evidence supporting no increases in prediction with imputation (Okbay, Beauchamp, et al., 2016; E. B. Ware et al., 2017). The PGSs were not adjusted for any covariates when being constructed (Ajnakina & Steptoe, 2022).

#### Exposures – polygenic scores

For mental health PGSs, we included two common mental health traits often captured in symptom scales even if at subclinical levels (i.e., PGS_depressive-symptoms_ and PGS_anxiety_) (Okbay, Baselmans, et al., 2016; Otowa et al., 2016) as well as PGSs for two severe mental illnesses, which are harder to identify at subclinical levels in population data and affect only relatively small numbers of individuals (i.e., PGS_bipolar-disorder_ and PGS_SCZ_) (Mullins et al., 2021; Trubetskoy et al., 2022). In addition, we also included PGS_wellbeing_ (Okbay, Baselmans, et al., 2016) to capture positive aspects of mental health, which, while related, are conceptually distinct from mental ill health (Keyes, 2005). For more detail on the PGSs see Supplementary B.

### Outcomes – social connection phenotypes

For social connection phenotypes, empirical evidence has found that structural, functional, and quality aspects of social connections are only weakly correlated and have independent relationships with various health outcomes (Holt-Lunstad, 2018; Holt-Lunstad et al., 2010, 2017). Therefore, we used separate measures of social connection for each domain. We looked at social isolation (structural aspect), loneliness and social support (functional aspects), and relationship strain (quality aspect) (Holt□ Lunstad, 2024). Social isolation is an objective measure of lack of contact with others due to actual physical distance or reduced social networks (Mansfield et al., 2019). Loneliness is broadly defined as the subjective experience of psychological distress due to a perceived disconnect in quantity and quality between one’s aspired and actual social connections (Hawkley & Cacioppo, 2010; Perlman & Peplau, 1998), while social support refers to the extent to which one has access to instrumental, informational, appraisal and emotional resources (Berkman & Krishna, 2014). Finally, relationship strain assesses the negativity within one’s social connections (Brooks & Dunkel Schetter, 2011).

Loneliness was measured using the UCLA 3-item Loneliness Scale, with higher scores indicating greater loneliness (Hughes et al., 2004). Social isolation was measured via the frequency of contact (meeting up and speaking on the phone) with children, family or friends, with higher scores indicating greater social isolation. Social support was measured via three items asking individuals how much each type of relationship (i.e., partner, children, family, friends) supported them, with higher scores indicating greater social support. Relationship strain was measured via three items asking individuals how much each type of relationship (i.e., partner, children, family, friends) strained them, with higher scores indicating greater relationship strain. For more details on these social measures, see Supplementary C.

### Covariates

Ten principal components (PCs) were added to the models to control for ancestral differences that could bias results (Price et al., 2006), and analyses were adjusted for chronological age (in years) and sex (male or female). It is recognised that binary measures of assigned sex at birth do not reflect all gender identities, but these were the only data available. Adjusting for these covariates only is standard practice in the genetics literature when using PGSs as exposures (Ajnakina et al., 2022; Kwong et al., 2021), whereby adjusting for phenotypic covariates could lead to bias in the models (e.g., phenotypic covariates could lie on the causal pathway from PGS exposure to phenotypic outcomes) (Akimova et al., 2021).

### Statistical analyses

To explore the associations between each of the five mental health PGSs and each of the four social connection phenotypic outcomes, we ran linear regressions in 20 separate analyses. Due to multiple testing, a Bonferroni correction was applied. As this is a conservative method, it was deemed sufficient for the significance level to be adjusted by the number of PGSs (p<0.01 (p=0.05/5 PGSs) (VanderWeele & Mathur, 2019). Each PGS was standardized to have a mean of 0 and a standard deviation (SD) of 1 to aid interpretability. All models were fitted using complete case analysis, excluding participants with missing data. All analyses controlled for age, sex, and 10 PCs. All analyses were conducted in STATA release 18.0 (STATA Corp LP, USA).

We also ran two sensitivity analyses. The first sensitivity analysis excluded individuals with a diagnosis closely related to the PGS exposure under investigation to see if this impacted the results (therefore sensitivity analyses were not applied to PGS_wellbeing_). In. Additionally, we ran a PGS for major depressive disorder (PGS_MDD_) to see if there were any differences in results compared to PGS_depressive-symptoms_. For details, see Supplementary D.

## Results

### Sample characteristics

The mean sample age was 65 years (SD=9.3) and 55% were female (Table 1). The mean score for social isolation was 5.0 (SD=1.8), 4.0 (SD=1.4) for loneliness, 7.7 (SD=2.4) for social support and 2.2 (SD=1.6) for relationship strain (see Table 1 for further characterisation of the sample).

**Table 1.** Sample characteristics.

|  | Mean (SD) |
| --- | --- |
| **\*Age** | 65.5, 9.3 |
| **\*Social isolation** | 5.0, 1.8 |
| **\*Loneliness** | 4.0, 1.4 |
| **\*Social support** | 7.7, 2.4 |
| **\*Relationship strain** | 2.2, 1.6 |
|  | Percentage (%) |
| **\*Gender** |  |
| *Female* | 54.6 |
| *Male* | 45.4 |
| **Wealth** |  |
| 1 - lowest quintile | 15.5 |
| 2 | 18.9 |
| 3 | 21.4 |
| 4 | 22.5 |
| 5 - highest quintile | 21.8 |
| **Education** |  |
| No qualification | 34.7 |
| NVQ1/CSE | 4.7 |
| NVQ2/GCE O-level | 18.4 |
| NVQ3/GCE A-level | 7.1 |
| Higher ed below degree | 13.3 |
| NVQ4/NVQ5/degree | 13.2 |
| Foreign/other | 8.6 |
| **Employment** |  |
| Not employed | 14.0 |
| Employed | 36.9 |
| Retired | 49.1 |
| **Number of people in household** |  |
| One | 22.4 |
| Two | 60.3 |
| Three | 11.7 |
| Four or more | 5.5 |
| **Number of group memberships** |  |
| None | 25.8 |
| One | 29.7 |
| Two | 22.4 |
| Three | 12.6 |
| Four or more | 9.5 |
| **Illness** |  |
| No | 44.8 |
| Yes, not limiting | 23.0 |
| Yes, limiting | 32.2 |

SD = standard deviation. Only the variables with asterisks were used in the analytical models (n=5,257). The sample sizes varied for some of the variables that are presented only for sample descriptives (wealth n=5,200, education n=5,254, group memberships n=3,522, illness, n=5,255).

### Main analyses

#### Social isolation

Higher PGS_wellbeing_ was associated with lower social isolation (B=-0.07, CI-95%=-0.12, -0.02, p=0.003) and higher PGS_anxiety_ was associated with higher social isolation (B=0.05, CI- 95%=0.00, 0.10, p=0.050). But there was no evidence of associations for PGS_depressive-symptoms,_ PGS_bipolar-disorder_ or PGS_SCZ_ (Figure 1, Supplementary Table S1). The association with PGS_anxiety_ did not remain after Bonferroni correction.

**Figure 1.**
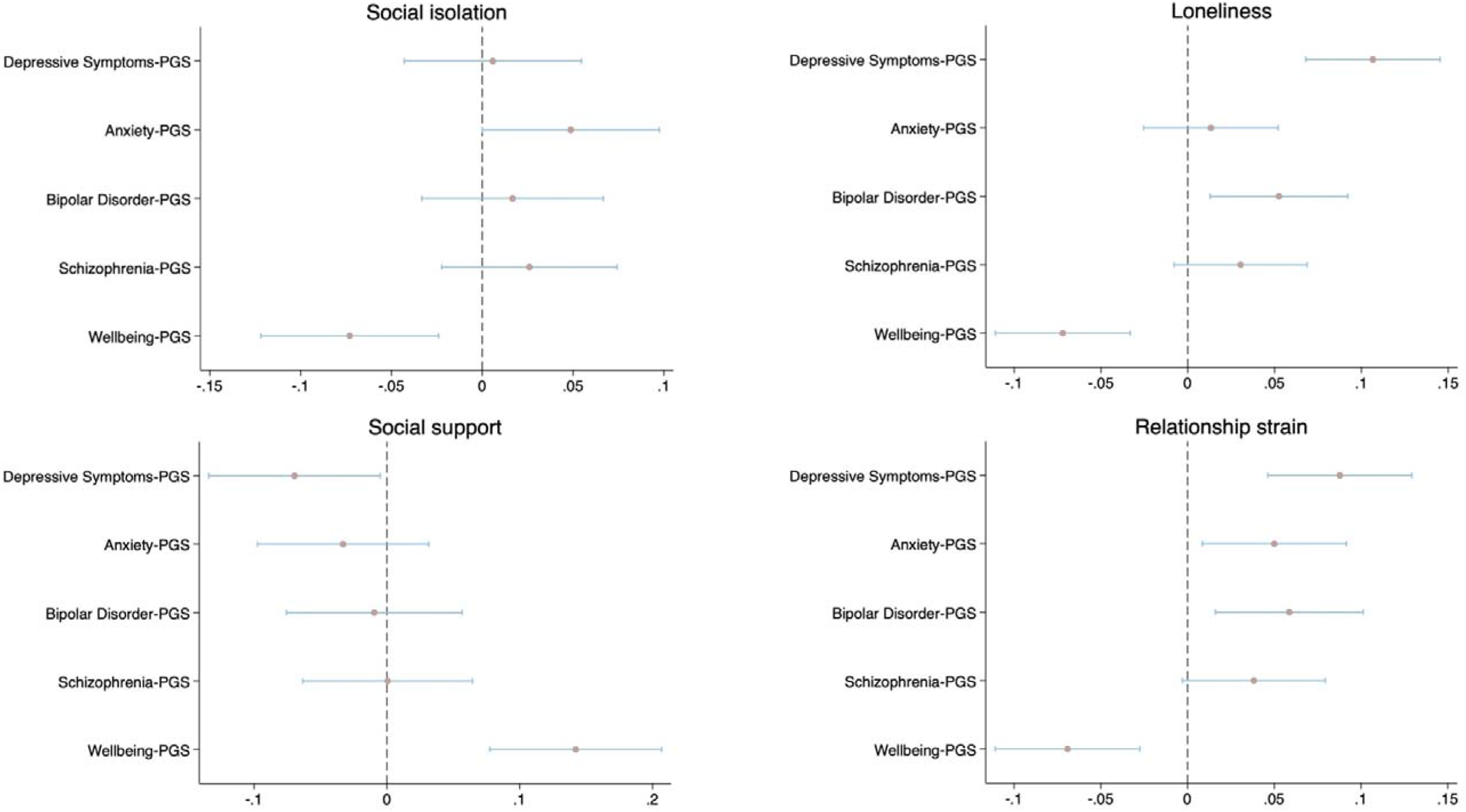
Associations between mental health PGSs and social connection phenotypes. PGS = polygenic score. The x-axis represents unstandardised beta estimates relating to a 1 standard deviation (SD) increase in PGS. All models controlled for age, sex and 10 principal components (PCs). Sample size is n=5,257.

#### Loneliness

Higher PGS_depressive-symptoms_ and PGS_bipolar-disorder_ were associated with higher loneliness (B=0.11, CI-95%=0.07, 0.15, p<.001; B=0.05, CI-95%=0.01, 0.09, p=0.009). In contrast, higher PGS_wellbeing_ was associated with lower loneliness (B=-0.07, CI-95%=-0.11, -0.03, p<.001). However, PGS_anxiety,_ and PGS_SCZ_ showed no associations with feeling lonely (Figure 1, Supplementary Table S1).

#### Social support

Higher PGS_depressive-symptoms_ was associated with lower perceived social support (B=-0.07, CI- 95%=-0.13, -0.01, p=0.035), while higher PGS_wellbeing_ was associated with higher social support (B=0.14, CI-95%=0.08, 0.21, p<.001). The other PGSs were not associated with social support (Figure 1, Supplementary Table S1). The association with PGS_depressive-symptoms_ did not remain after Bonferroni correction.

#### Relationship strain

Higher PGS_depressive-symptoms,_ PGS_anxiety,_ and PGS_bipolar-disorder_ were associated with higher perceived relationship strain (B=0.09, CI-95%=0.05, 0.13, p<.001; B=0.05, CI-95%=0.01, 0.09, p=0.018; B=0.06, CI-95%=0.02, 0.10, p=0.007). Whereas higher PGS_wellbeing_ was associated with lower relationship strain (B -0.07, CI-95%=-0.11, -0.03, p=0.001). Relationship strain was not associated with PGS_SCZ_ (Figure 1, Supplementary Table S1). The association with PGS_anxiety_ did not remain after Bonferroni correction.

### Sensitivity analyses

The pattern of associations when individuals with a related diagnosed mental health condition were removed from the analyses were fairly homogenous with the main findings (Supplementary Table S2). However, the association between PGS_depressive-symptoms_ and social support was no longer evident (B=-0.06, CI-95%=-0.13, 0.00, p=0.058). Additionally, PGS_MDD_ showed the same pattern of results as PGS_depressive-symptoms_ (Supplementary Table S3).

## Discussion

This paper identified unique associations between different mental health PGSs and different aspects of social connection phenotypes in older adults. A genetic propensity to higher wellbeing was related to all aspects of social connections. A higher genetic propensity to depressive symptoms was related only to functional and quality aspects (loneliness, social support and relationship strain). A higher genetic propensity to anxiety was related just to structural and quality aspects (social isolation and relationship strain). A higher genetic propensity to bipolar disorder was only related to one measure of functional aspects (loneliness), as well as quality aspects (relationship strain). A genetic propensity to schizophrenia was not related to any aspects of social connections. Findings were not materially altered in sensitivity analyses. Associations between mental health PGSs and social connections have been evidenced in previous studies (Abdellaoui et al., 2018; Richardson et al., 2019). However, our findings advance such work by applying an established theoretical framework for conceptualising social connections (Holt□ Lunstad, 2024) and identifying more nuanced relationships than previously known.

The associations between higher genetic propensity to depression, bipolar and anxiety and poorer social connections typically aligns with the phenotypic literature. Individuals with higher phenotypic depressive symptoms tend to have high loneliness and strain in their connections and low perceived social support (Ali et al., 2022; Dahlberg et al., 2021; Turner et al., 2022), as do individuals with bipolar disorder (Eidelman et al., 2012; Koenders et al., 2015; Pike et al., 2024). Individuals with phenotypic anxiety also typically have poorer social connections (e.g., more conflict in their relationships) (Leach & Butterworth, 2020). Notably, these conditions are linked to poorer social functioning and impaired social cognition, such as social avoidance and withdrawal, not recognising other’s emotions, and negative evaluations of social stimuli (Gillissie et al., 2022; Saris et al., 2017; Weightman et al., 2014), potentially explaining why associations were strongest for more subjective measures relating to function and quality aspects of social connections, but also why higher genetic propensity to anxiety (potentially if anxiety is manifesting as social anxiety) was associated with social isolation too. In contrast, a higher genetic propensity to wellbeing was associated with better social connections for all four outcomes. Again, this mirrors findings in the phenotypic literature: individuals with higher phenotypic mental wellbeing tend to have low loneliness, social isolation, and strain in their relationships, as well as high perceived social support (Ermer & Proulx, 2022; Shankar et al., 2015; Siedlecki et al., 2014; VanderWeele et al., 2012). PGS_wellbeing_ was derived using measures of positive affect and life satisfaction, which are evaluative and experienced forms of hedonic wellbeing, respectively (Stone & Mackie, 2013). So, some of the observed associations (especially for subjective aspects of social connections) could indicate more positive perspectives on one’s life. Additionally, individuals with higher wellbeing may be more proactive in engaging socially with others, potentially leading to stronger social networks and reduced social isolation.

There were no findings for PGS_SCZ_, which does not align with the phenotypic literature. SCZ also affects social cognition processes (e.g., perception of social cues and experience sharing) that can negatively impact social connections (Dong et al., 2019; Green et al., 2015). High-risk individuals have reduced social networks, higher loneliness, less perceived social support, and poorer quality connections (Robustelli et al., 2017). This divergence may be because our analyses examined the role of genetic propensity to SCZ within a general population sample rather than a clinical sample. There could also be a complex interplay between SCZ and specific aspects of social connections when considering genetics. Indeed, Mendelian Randomization has found a bidirectional causal effect between SCZ and loneliness (functional aspect), a unidirectional effect of ability to confide in others (functional aspect) on SCZ and SCZ on number of people in household (structural aspect), but no causal effect between the number of family/friends visits (structural aspect) and SCZ (Andreu-Bernabeu et al., 2022). However, the PGS_SCZ_ findings also do not align with existing PGS-phenotype literature, where it has been found to be associated with loneliness (Socrates et al., 2021).

In addition to the behavioural mechanisms just discussed, it is relevant to consider other mechanisms for how mental health PGSs could influence social behaviours. Phenotypic mental health (e.g., depression, bipolar disorder) and mental resilience have been linked to inflammatory processes and the modulation of stress systems (Maletic & Raison, 2014; Ryan & Ryznar, 2022; Strawbridge et al., 2018). For example, greater mental wellbeing (i.e., living a meaningful life) has been associated with more favourable inflammatory profiles in older adults (Steptoe & Fancourt, 2019). Inflammation can translate into social environments by inducing sickness behaviours, such as altered sociability, including both social withdrawal and social approach (i.e., seeking supportive connections) (Charles et al., 2020; Devlin et al., 2022; Muscatell & Inagaki, 2021). Indeed, higher loneliness may be bidirectionally associated with higher inflammation (Gao et al., 2024), lower social support and higher relationship strain are related to greater cortisol levels (Iob et al., 2018), and lower multi- system inflammation and stress profiles (i.e., measured via allostatic load) is related to greater social activity via leisure engagement (Wang et al., 2021) and better functional aspects of social connections, such as higher spousal support (Brooks et al., 2014). It is notable that in the depressive symptoms, wellbeing, and bipolar disorder GWASs, analyses to identify the functions of identified genetic variants found enrichment in tissue and biological processes implicated in inflammatory and stress systems (Okbay, Baselmans, et al., 2016; Stahl et al., 2019).

However, the exact mechanisms underpinning associations between mental health PGSs and social connection phenotypes are complex to unpack (Socrates et al., 2021). Genetic variants may exert themselves directly through biological pathways via pleiotropy (where the same genetic variants influence multiple traits) (Hackinger & Zeggini, 2017). This could be horizontal pleiotropy, where genetic variants associated with mental health and social connections traits are the same or localised in the same region and may independently influence both, or vertical pleiotropy, where genetic variants associated with mental health traits may influence social connections through a mediating pathway. Genetic variants for mental health traits could also exert themselves indirectly by shaping social environments. Individuals more genetically prone to higher wellbeing may select more socially fulfilling environments. Lastly, there is genetic overlap (measured via genetic correlations) between the traits of depressive symptoms, anxiety, bipolar disorder, and wellbeing (Okbay, Baselmans, et al., 2016). This could explain why these specific mental health traits show patterns with social connection phenotypes. However, SCZ is also genetically correlated to these traits, depression, yet we do not see the same pattern of association (Anttila et al., 2018; Okbay, Baselmans, et al., 2016). This suggests there are distinct associations between genetic propensity to depressive symptoms, anxiety, bipolar disorder, and wellbeing with different aspects of social connections (i.e., structural, functional, and quality).

The paper has multiple strengths. It addresses temporality concerns, a common issue in observational phenotypic research, by assessing how mental health PGSs are associated with social connection phenotypes in cross-sectional analyses. It explores how genetic propensity for mental health traits not generally measured in population samples or not expressed phenotypically (such as bipolar disorder) influence social connection behaviours. We carefully situated our social connection phenotypes and interpretation of findings within social theory, which has been lacking in existing genetics studies. However, due to genetic data availability, we could only include individuals of White European ancestry in our analyses, which is a substantial concern in health research and affects the generalisability of findings (Martin et al., 2019). This lends to consideration of important ethical and societal implications when using genetic data (i.e., understanding that genetics are not deterministic) (Harden & Koellinger, 2020; Martschenko, 2022). Indeed, the genetic variants used in these PGSs only explain small variances of their respective phenotypic trait (Visscher et al., 2017), and both phenotypic social connections and phenotypic mental health traits are influenced by more than just genetic factors (i.e., exogenous social factors). Findings also need to be taken in light of how the PGSs and social connection phenotypes were derived and measured (e.g., relating to the GWAS sample size, the operalisation of the phenotypic measurement used in the GWAS, and our social connection constructs) (Dudbridge, 2013; Escott-Price & Hardy, 2022). However, adding strength to our choices, we used an established theoretical framework (Holt□ Lunstad, 2024) and well-validated or frequently used indices for our social constructs.

Overall, we demonstrate differential associations between mental health PGSs and distinct aspects of social connections, indicating a nuanced picture. Future work is encouraged to explore how these mental health PGSs and other mental health traits (e.g. obsessive-compulsive disorder) are associated with trajectories of social connections over time, as well as potential moderation effects (e.g. by socio-economic positions) and the consistency of findings in different populations. Our findings suggest that genetics may influence social behaviours that could, in turn, act as risk-enhancing or risk-reducing factors for the experience of phenotypic symptoms of mental ill-health. This furthers the importance of interventions looking to connect individuals at risk of mental illness as well as those showing symptoms with social activities.

## Supporting information

Finn et al. 2024 - Supplement

## Data Availability

The data are freely available through the UK data services and can be accessed here: https://discover.ukdataservice.ac.uk

## Acknowledgements

This work is supported by UK Research and Innovation [MR/Y01068X/1] and this work was developed from SF’s PhD work, which was supported by the ESRC-BBSRC Soc-B Centre for Doctoral Training (ES/P000347/1).

