## Supplementary material for "Genetic propensity to mental health traits and their associations with social connection phenotypes: evidence from the English Longitudinal Study of Ageing": Finn et al. 2024 - Supplement

**Supplementary A.** Exposures: Polygenic scores.

$PGS_{depressive\text{-}symptoms}$ was created from a GWAS meta-analysis of 180,866 individuals of European ancestry (Okbay, Baselmans, et al., 2016). Depression was phenotyped using continuous measures assessing depressive symptoms (i.e., two items from the Patient Health Questionnaire) and case-control data of major depressive disorder (MDD) (i.e., using ICD-9 or DSM classifications).

$PGS_{anxiety}$ was created from a GWAS meta-analysis of over 18,000 individuals of European ancestry (Otowa et al., 2016). A quantitative factor score was derived across disorders of generalised anxiety disorder (GAD), panic disorder, agoraphobia, social phobia, and specific phobia from DSM-based criteria.

$\mathrm{PGS}_{\mathrm{bipolar\text{-}disorder}}$ was created from a GWAS meta-analysis of 41,917 cases and 371,549 controls of European ancestry (Mullins et al., 2021). Lifetime diagnosis of bipolar disorder using diagnostic criteria (DSM-IV, ICD-9 or ICD-10) were included as cases.

$\mathrm{PGS}_{\mathrm{SCZ}}$ used data from a trans-ancestry GWAS meta-analysis of 76,755 individuals with a SCZ diagnosis and 243,649 controls (Trubetskoy et al., 2022).

$PGS_{wellbeing}$ used data from a GWAS meta-analysis of 298,420 individuals of European ancestry (Okbay, Baselmans, et al., 2016). Subjective wellbeing was phenotyped using measures of life satisfaction and positive affect).

**Supplementary B.** Outcomes: social connection measures.

***Loneliness*** was measured using the UCLA 3-item Loneliness Scale, a validated tool for population surveys (Hughes et al., 2004). The three items were rated on a 3-point scale (1=Hardly ever/never, 2=Some of the time, 3=Often) and were summed to create a score ranging from 3-9, with higher scores indicating higher levels of loneliness.

***Social isolation*** was measured via the frequency of contact (meeting up and speaking on the phone) with children, family or friends. This was measured on a 4-point scale (0=Three or more times a week, 1= Once or twice a week, 2=Once or twice a month or every few months, 3=Never or rarely). The scores for the three relationships were combined, with total scores ranging from 0-9 and higher scores indicating higher social isolation. As this is an objective measure of social connections, those who were unable to score for some of these components (e.g. because they have no children) were still included in the score (scored as 3=Never or rarely). Therefore, a total score was provided even if individuals did not have all three relationships. In the final analytical sample, n=4,904 had all three relationship types; n=313 had two relationship types, and n=40 had one relationship type.

***Social support*** was measured via three items asking individuals how much each type of relationship (i.e., partner, children, family, friends) supported them. This included: i) How much do they really understand the way you feel about things, ii) How much can you rely on them if you have a serious problem?=, and iii) How much can you open up to them if you need to talk about your worries. This was measured on a 4-point scale (1=Not at all, 2=A little, 2=Some of the time, 3=A lot). A combined social support score was constructed by summing average scores across the three items for all four relationships, ranging from 0-12, with higher scores indicating greater perceived social support. This meant that individuals with one or all four relationships were included in the score. In the final analytical sample, n=3,045 had all four relationship types, n=1,532 had three relationship types, n=582 had two relationship types, and n=98 had one relationship type. In the self-report questionnaire, individuals did not answer this question if the relationship did not exist. Unlike the social isolation measure, which is an objective construct, individuals without sources of support from not having specific relationships were not included in this score. This is because social support aimed to measure subjective perceptions of existing relationships.

***Relationship strain*** was measured via three items asking individuals how much each type of relationship (i.e., partner, children, family, friends) gave them strain. This included: i) How much do they criticise you, ii) How much do they let you down when you are counting on them, and iii) How much do they get on your nerves. This was measured on a 4-point scale (1=Not at all, 2=A little, 2=Some of the time, 3=A lot). A combined relationship strain score was constructed by summing average scores across the three items for all four relationships ranging from 0-12, with higher scores indicating higher relationship strain. This meant that individuals with one or all four relationships were included. In the final analytical sample, n=2,928 had all four relationship types, n=1,552 had three relationship types, n=641 had two relationship types, and n=136 had one relationship type. In the self-report questionnaire, individuals did not answer this question if the relationship did not exist. Therefore, individuals without sources of strain from not having specific relationships were not included in this score. This is because relationship strain aimed to measure subjective perceptions of existing relationships.

**Supplementary C.** Details of sensitivity analyses.

Sensitivity analysis 1.

Individuals were asked if they had ever been told they have an emotional, nervous or psychiatric problem by a doctor and if so what type. Prompts were given listing hallucinations, anxiety, depression, emotional problems, psychosis, mood swings, manic depression, or something else, therefore, individuals could identify with more than one. Individuals who had mentioned depression or anxiety were identified as depression and anxiety diagnoses, respectively. Individuals were not asked about having a diagnosis of bipolar disorder specifically, however, mentions of mood swings or manic depression identified bipolar disorder diagnoses. Individuals were not asked about having a diagnosis of SCZ specifically, however, mentions of hallucinations or psychosis identified SCZ diagnoses.

Sensitivity analysis 2.

$PGS_{MDD}$ used data from a GWAS meta-analysis of 135,458 cases and 344,901 controls of European ancestry, where lifetime MDD was measured via diagnostic criteria (e.g., DSM and ICD).[1]

[1] Wray, N. R., Ripke, S., Mattheisen, M., Trzaskowski, M., Byrne, E. M., Abdellaoui, A., ... & Viktorin, A. (2018). Genome-wide association analyses identify 44 risk variants and refine the genetic architecture of major depression. *Nature genetics*, *50*(5), 668-681.

Finn et al. (2024). Genetic propensity to mental health traits and their associations with social connection phenotypes: evidence from the English Longitudinal Study of Ageing.

**Table S1.** Associations between mental health PGSs and social connection phenotypes.

| | Social isolation | | | Loneliness | | | Social support | | | Relationship strain | | |
|---|---|---|---|---|---|---|---|---|---|---|---|---|
| | B | CI-95% | p-value | B | CI-95% | p-value | B | CI-95% | p-value | B | CI-95% | p-value |
| **$PGS_{depressive\text{-}symptoms}$** | 0.01 | -0.04, 0.05 | 0.816 | **0.11** | **0.07, 0.15** | **<.001** | **-0.07** | **-0.13, -0.01** | **0.035** | **0.09** | **0.05, 0.13** | **<.001** |
| **$PGS_{anxiety}$** | **0.05** | **0.00, 0.10** | **0.050** | 0.01 | -0.03, 0.05 | 0.500 | -0.03 | -0.10, 0.03 | 0.317 | **0.05** | **0.01, 0.09** | **0.018** |
| **$PGS_{bipolar\text{-}disorder}$** | 0.02 | -0.03, 0.07 | 0.512 | **0.05** | **0.01, 0.09** | **0.009** | -0.01 | -0.08, 0.06 | 0.778 | **0.06** | **0.02, 0.10** | **0.007** |
| **$PGS_{SCZ}$** | 0.03 | -0.02, 0.07 | 0.292 | 0.03 | -0.01, 0.07 | 0.119 | 0.00 | -0.06, 0.06 | 0.987 | 0.04 | -0.00, 0.08 | 0.069 |
| **$PGS_{wellbeing}$** | **-0.07** | **-0.12, -0.02** | **0.003** | **-0.07** | **-0.11, -0.03** | **<.001** | **0.14** | **0.08, 0.21** | **<.001** | **-0.07** | **-0.11, -0.03** | **0.001** |

PGS = polygenic score. B = unstandardised beta estimates relating to a 1 standard deviation (SD) increase in PGS. CI-95% = 95% confidence intervals. All models controlled for age, sex and 10 principal components (PCs). Bold estimates indicate evidence of association. Sample size is n=5,257.

**Table S2.** Sensitivity analyses: associations between mental health PGSs and social connection phenotypes when removing individuals with a diagnosed mental health condition.

| | **Social isolation** | | | **Loneliness** | | | **Social support** | | | **Relationship strain** | | |
|---|---|---|---|---|---|---|---|---|---|---|---|---|
| | B | CI-95% | p-value | B | CI-95% | p-value | B | CI-95% | p-value | B | CI-95% | p-value |
| **PGS**$_{depressive\text{-}symptoms}$ | 0.00 | -0.05, 0.05 | 0.939 | 0.10 | **0.06, 0.14** | **<.001** | -0.06 | -0.13, 0.00 | 0.058 | 0.08 | **0.04, 0.12** | **<.001** |
| **PGS**$_{anxiety}$ | 0.05 | -0.00, 0.10 | 0.051 | 0.01 | -0.03, 0.05 | 0.715 | -0.02 | -0.09, 0.04 | 0.459 | 0.05 | **0.01, 0.09** | **0.016** |
| **PGS**$_{bipolar\text{-}disorder}$ | 0.02 | -0.03, 0.07 | 0.503 | 0.05 | **0.01, 0.09** | **0.015** | -0.01 | -0.07, 0.06 | 0.825 | 0.06 | **0.02, 0.10** | **0.006** |
| **PGS**$_{SCZ}$ | 0.03 | -0.02, 0.07 | 0.298 | 0.03 | -0.01, 0.07 | 0.115 | 0.00 | -0.06, 0.06 | 0.986 | 0.04 | -0.00, 0.08 | 0.069 |

PGS = polygenic score. B = unstandardised beta estimates relating to a 1 standard deviation (SD) increase in PGS. CI-95% = 95% confidence intervals. All models controlled for age, sex and 10 principal components (PCs). Bold estimates indicate evidence of association. The sample sizes differed slightly (**PGS**$_{depressive\text{-}symptoms}$ n=5,172, PGS$_{anxiety}$ n=5,185, **PGS**$_{bipolar\text{-}disorder}$ n=5,240, **PGS**$_{SCZ}$ n=5,252.

Finn et al. (2024). Genetic propensity to mental health traits and their associations with social connection phenotypes: evidence from the English Longitudinal Study of Ageing.

**Table S3.** Sensitivity analyses: $PGS_{MDD}$ associations with social connection phenotypes.

| | **Social isolation** | | | **Loneliness** | | | **Social support** | | | **Relationship strain** | | |
|---|---|---|---|---|---|---|---|---|---|---|---|---|
| | B | CI-95% | p-value | B | CI-95% | p-value | B | CI-95% | p-value | B | CI-95% | p-value |
| **$PGS_{MDD}$** | 0.01 | -0.04, 0.06 | 0.767 | **0.12** | **0.08, 0.16** | **<.001** | **-0.13** | **-0.19, -0.06** | **<.001** | **0.10** | **0.06, 0.14** | **<.001** |

PGS = polygenic score. MDD = major depressive disorder. B = unstandardised beta estimates relating to a 1 standard deviation (SD) increase in PGS. CI-95% = 95% confidence intervals. All models controlled for age, sex and 10 principal components (PCs). Bold estimates indicate evidence of association. Sample size is n=5,257.
